# Personality traits are consistently associated with blood mitochondrial DNA copy number estimated from genome sequences in two genetic cohort studies

**DOI:** 10.1101/2022.06.04.22275970

**Authors:** Richard F. Oppong, Antonio Terracciano, Martin Picard, Yong Qian, Thomas J. Butler, Toshiko Tanaka, Ann Zenobia Moore, Eleanor M. Simonsick, Krista Opsahl-Ong, Christopher Coletta, Angelina R. Sutin, Myriam Gorospe, Susan M. Resnick, Francesco Cucca, Sonja W. Scholz, Bryan J. Traynor, David Schlessinger, Luigi Ferrucci, Jun Ding

## Abstract

**Background:** Mitochondrial DNA copy number (mtDNAcn) in tissues and blood can be altered in conditions like diabetes and major depression and may play a role in aging and longevity. However, little is known about the association between mtDNAcn and personality traits linked to emotional states, metabolic health, and longevity. This study tests the hypothesis that blood mtDNAcn is related to personality traits and mediates the association between personality and mortality.

**Methods:** We assessed the big five personality domains and facets using the Revised NEO Personality Inventory (NEO-PI-R), assessed depressive symptoms with the Center for Epidemiologic Studies Depression Scale (CES-D), estimated mtDNAcn levels from whole-genome sequencing, and tracked mortality in participants from the Baltimore Longitudinal Study of Aging. Results were replicated in the SardiNIA Project.

**Results:** We found that mtDNAcn was negatively associated with the Neuroticism domain and its facets and positively associated with facets from the other four domains. The direction and size of the effects were replicated in the SardiNIA cohort and were robust to adjustment for potential confounders in both samples. Consistent with the Neuroticism finding, higher depressive symptoms were associated with lower mtDNAcn. Finally, mtDNAcn mediated the association between personality and mortality risk.

**Conclusions:** To our knowledge, this is the first study to show a replicable association between mtDNAcn and personality. Furthermore, the results support our hypothesis that mtDNAcn is a biomarker of the biological process that explains part of the association between personality and mortality.

## Introduction

Mitochondria are cellular organelles essential for the life of eukaryotic cells (1). They produce adenosine 5’-triphosphate (ATP), the main energetic currency for biological function, by oxidative phosphorylation (OXPHOS) of a number of substrates (2). Mitochondria harbor their own circular DNA molecules (mtDNA), which lack histone proteins and bear genes lacking introns (2). The proximity of mtDNA to OXPHOS production and its lack of histone protection makes it more susceptible to ROS damage, threatening healthy cellular function (3). The number of copies of mtDNA (mtDNAcn) in tissues and blood leukocytes is considered an imperfect proxy measure of mitochondrial mass that may reflect energetic resilience or hematopoiesis (4).

Consequently, reduced mtDNAcn in peripheral blood has been associated with various health outcomes, including renal cell carcinoma (5), hypertension (6), neurodegenerative disease (7), and diabetes-related conditions, including insulin sensitivity (8, 9), fasting plasma glucose level (12), and diabetic nephropathy (10). High levels of mtDNAcn in tissue and the blood can also occur in conditions of stress and illness such as diabetes (13) and peripheral artery disease (14), suggesting that in some situations of stress, mtDNAcn can be elevated to compensate for low cellular energy (4, 15) and stress adaptation (16). For major depression, however, the findings are mixed, with some studies reporting that mtDNAcn has a positive (17), negative (18), or null association with depression (19–23).

Since specific personality traits affect individuals’ susceptibility to environmental stress (24) and the development of pathology and premature death, we hypothesized that blood mtDNAcn is a biomarker of stress burden and might mediate the association between personality traits and excess mortality (25). To test this hypothesis, we tested the relationship between personality and levels of whole-blood mtDNAcn estimated by whole-genome sequencing and whether mtDNAcn mediated the relation between personality and mortality. To ensure replicability and robustness, we tested these associations in two cohorts, one from the US and the other from Italy.

To assess personality, we used participants’ profiles of the *big five* domains, including the six facets of each domain, measured with the Revised NEO Personality Inventory (NEO-PI-R), a personality measure validated extensively in multiple populations and languages (26). Personality traits measured with the NEO-PI-R have been associated with stressors such as anxiety and depression (27, 28), cognitive decline (29, 30), metabolic dysfunction (31, 32), energy levels (33), and physical illness (34). Facets of personality have also been associated with mortality risk (25). Compared with behavioral pathways to mortality risk (35), there has been less attention on the physiological mechanisms that explain the well-known association between personality and mortality risk. As mentioned above, lower mtDNAcn has been associated with both stressful situations and higher mortality (36). We thus address the critical novel question by testing whether mtDNAcn mediates the association between personality traits and mortality risk.

The goal of this study was to investigate the association between blood mtDNAcn and personality traits and facets assessed by the NEO-PI-R in two epidemiological studies: the Baltimore Longitudinal Study of Aging (BLSA) (37, 38) and the SardiNIA study (39). Using mediation analysis, we further tested the hypothesis that mtDNAcn is a biomarker in the biological process that connects personality characteristics with mortality.

## Results

### Relationship of sex, age, and white blood cell parameters with mtDNAcn

In both cohorts, mtDNAcn was negatively associated with age and male biological sex (Table S1). Furthermore, higher mtDNAcn was associated with lower WBC count and lower neutrophil percentage and was also associated with higher platelet count, lymphocyte percentage, eosinophil percentage, monocyte percentage, and basophil percentage (Table S1), consistent with previous findings (4). After adjusting for the effect of sex, sequence coverage, platelet count, and white blood cell parameters, there remained a significant, albeit modest, inverse association of mtDNAcn with age in both study cohorts (Figure 1 and Table S1). For the BLSA cohort, on average, every one-year increase in age was associated with a 0.014 standard deviation decrease in mtDNAcn (about 3.89% decrease in mtDNAcn per decade of life, *p*-value = 2.5 × 10^−5^), while in the SardiNIA cohort, there was a 0.0065 standard deviation decline in mtDNAcn for every year increase in age (about 1.36% decrease in mtDNAcn per decade of life, *p*-value =0.028). After adjusting for all the other covariates, women still had a significantly higher mtDNAcn than men (Figure 1 and Table S1).

**Figure 1.**
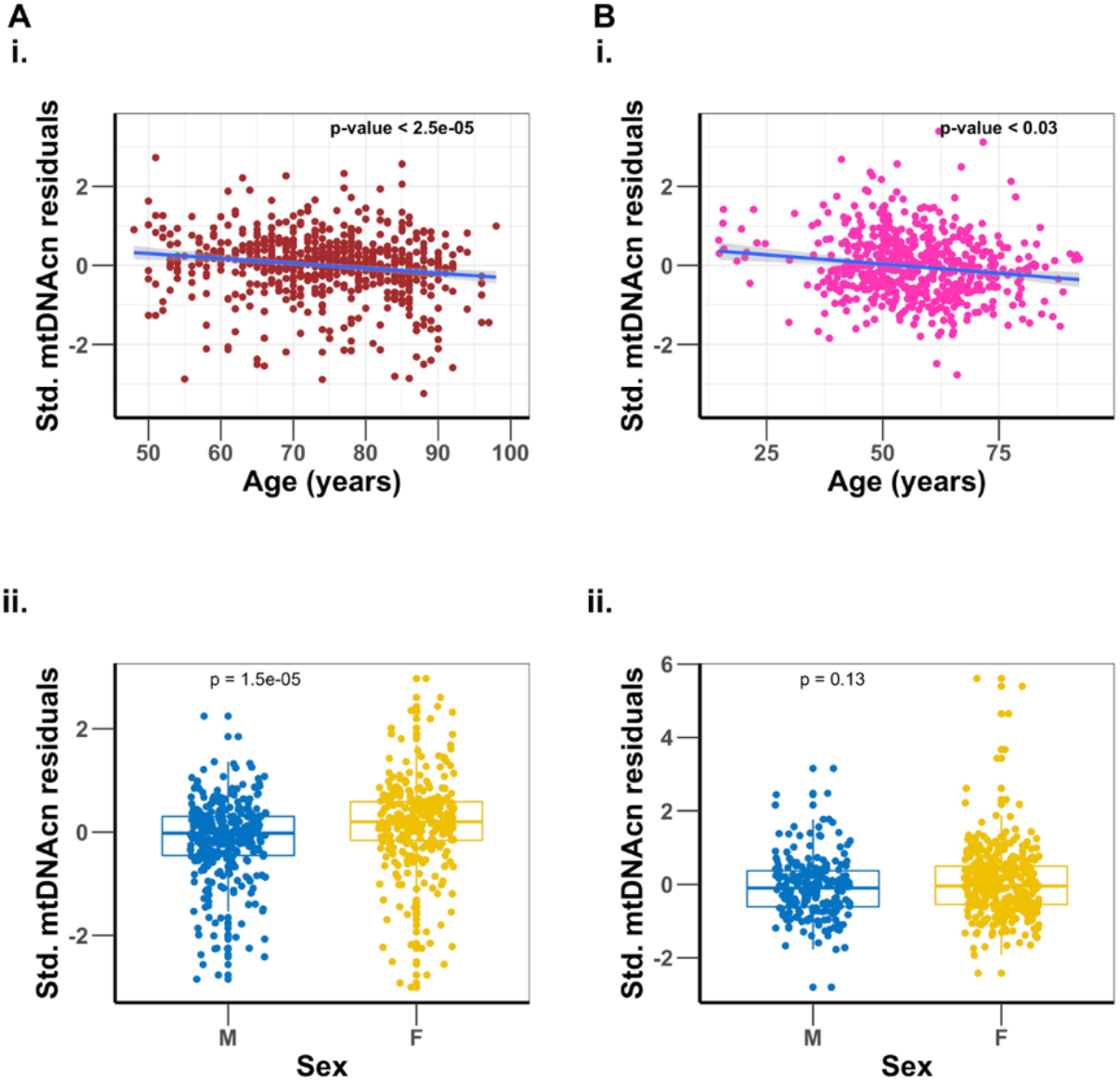
Association of mtDNAcn with age, and sex differences. Panel A is the BLSA cohort and panel B is the SardiNIA cohort. Plots i. are regression of mtDNAcn residuals on age after adjusting for sex, sequence coverage, platelets count and white blood cell parameters. Plots ii. are comparisons of mtDNAcn residuals after adjusting for age, sequence coverage, platelets count and white blood cell parameters between males (M) and females (F), with t-test p-values shown.

### Association of personality traits with mtDNA copy number

To test the association between NEO PI-R traits and mtDNAcn in each study cohort, we regressed mtDNAcn values on each personality trait, adjusting for age, sex, sequence coverage, platelet count, and white blood cell parameters. In both samples, mtDNAcn was associated with lower Neuroticism and higher Extraversion, Openness, Agreeableness, and Conscientiousness (Figure 2). We then performed a random-effects meta-analysis and used an FDR-corrected p-value threshold of 0.01. Among the 35 personality traits (five domains, 30 facets) tested, nine (five facets in the Neuroticism domain and four from Extraversion, Agreeableness, and Conscientiousness) were associated significantly with mtDNAcn even after the stringent FDR correction. Table 2 shows the individual study and meta-analysis results for the Neuroticism domain, while Table S2 provides detailed results for all 35 NEO PI-R traits.

**Table 1.**
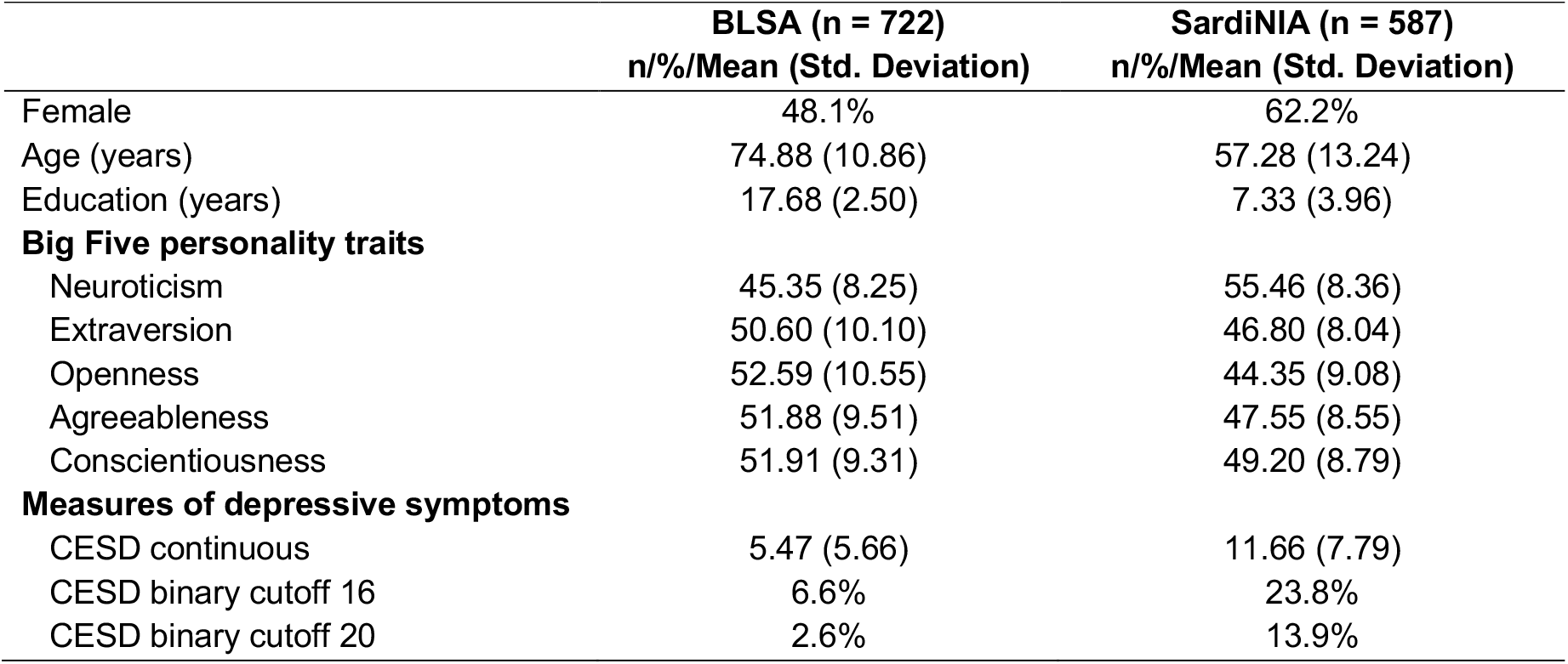
Demography of study cohorts.

**Table 2.**
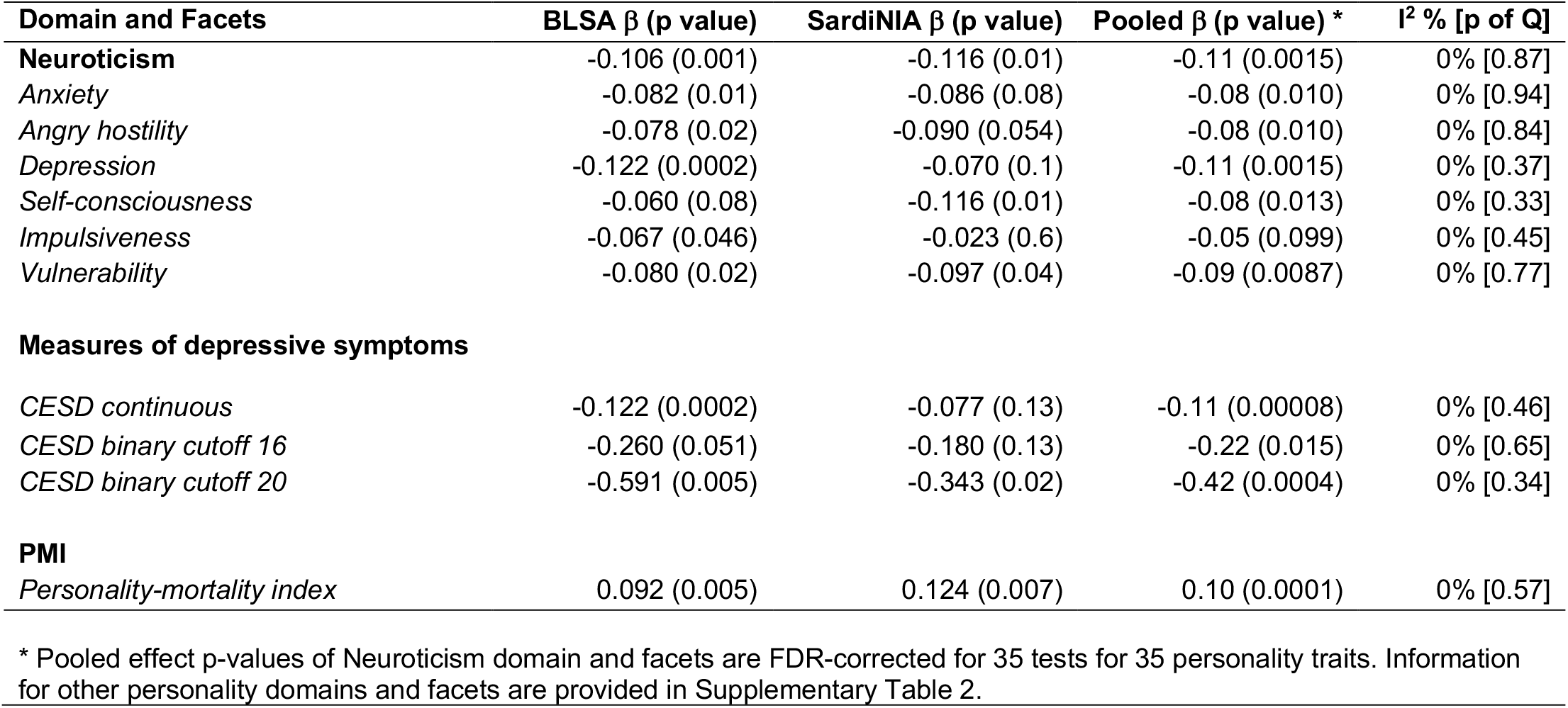
Association of Neuroticism domain with mtDNAcn.

**Figure 2.**
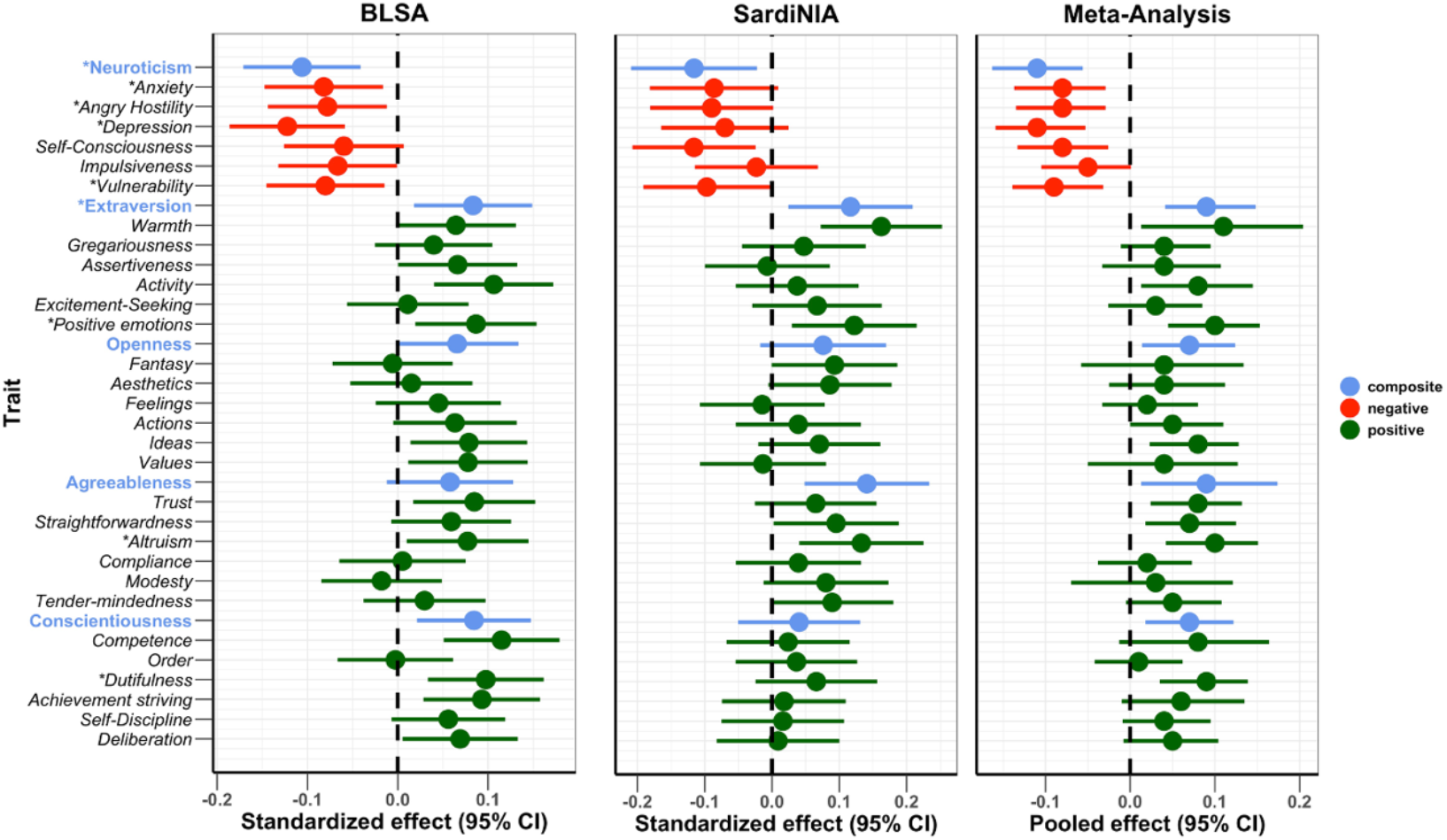
Association of NEO PI-R traits with mtDNAcn. Standardized/ Pooled effects with 95% CI of NEO PI-R traits after adjusting for the effects of age, sex, sequence coverage, platelets count and white blood cell parameters. Color coding is green for positive personality types, red for negative personality types and blue for the composite scores for the Big five NEO PI-R domains. * *Traits with FDR-corrected meta-analysis p-values <= 0*.*01*

The Neuroticism domain showed the most significant associations with mtDNAcn. The overall Neuroticism domain and four out of six facets were associated significantly (FDR p-value <= 0.01) with mtDNAcn (Table 2). The four facets were anxiety, angry hostility, depression, and vulnerability. Of note, facet self-consciousness was associated with mtDNAcn with an FDR-corrected p-value of 0.013. Notably, all neuroticism traits were associated negatively with mtDNAcn, with higher trait values corresponding to lower mtDNAcn values. Furthermore, the higher-order neuroticism domain was the personality trait associated with mtDNAcn with the most significant p-value (FDR p-value = 0.0015). The Neuroticism domain also showed highly consistent associations between the two study cohorts, with *I*^*2*^ (a measure of inconsistency among individual studies) equal to 0 for Neuroticism and its facets. Figure S1 shows the effect sizes for the Neuroticism domain and facets.

The overall Extraversion domain, the *positive emotions* facet in the Extraversion domain, the *altruism* facet in the Agreeableness domain, and the *dutifulness* facet in the Conscientiousness domain were associated with mtDNAcn with FDR-corrected p-values less than 0.01 (Table S2). These are all positive associations, with higher trait values corresponding to higher mtDNAcn values. No facets in the Openness domain were significantly associated with mtDNAcn.

Figure S2 compares the standardized effect sizes of the NEO PI-R traits between the two study cohorts. The results show remarkable concordance between the two study cohorts, with the data points clustering close to the diagonal line. In a sensitivity analysis evaluating the robustness of these findings to covariate adjustments, we compared the standardized effect sizes of the personality traits using models with and without adjustment for platelet count and white blood cell parameters (Fig S3). As most of the points were closely aligned to the diagonal line, we concluded that adjusting for platelet count and white blood cell parameters did not significantly impact the results.

### Association of CES-D with mtDNAcn

In both cohorts, we also regressed mtDNAcn on both continuous and binary CES-D values, adjusting for the effect of other covariates. Higher CES-D scores, measured as a continuous variable, were associated with lower mtDNAcn. Participants with depressive symptoms that exceeded the CES-D cutoff of either 16 or 20, had 6.06% or 11.57% lower mtDNAcn than those with no depressive symptoms, respectively (Table 2). Remarkably, in both cohorts, the effect sizes almost doubled when moving from moderate depressive symptoms (CES-D binary cutoff 16) to significant depressive symptoms (CES-D binary cutoff 20). The random-effects meta-analysis supported both the magnitude and direction of the effects with highly significant association *p-* values. There was also high consistency across the two cohorts for the continuous and binary CES-D traits, with *I*^*2*^ equal to 0.

### Personality, mtDNAcn, and mortality risk

We further studied the relationship between mtDNAcn and personality facets known to be associated with mortality risk. Figure S4 compares the mtDNAcn amongst four distinct personality types (see Methods for details) based on the *vulnerability* facet of Neuroticism and *self-discipline* facet of Conscientiousness, which were the two individual facets most strongly associated with mortality (25). Comparing the mtDNAcn between the four personality types in the BLSA cohort, we observed that the personality type with the perceived highest mortality risk (HVLD: <u>h</u>igh <u>v</u>ulnerability low self-<u>d</u>iscipline) had the lowest mtDNAcn amongst the four personality types. The HVLD personality type had a mean mtDNAcn value 0.23 standard deviations lower than the type with a perceived lowest mortality risk (LVHD: low <u>v</u>ulnerability <u>h</u>igh <u>s</u>elf-discipline) (*p*-value <0.0065), indirectly supporting an association between blood mtDNAcn and mortality risk. A more detailed description and interpretation of the results can be found in the Supplementary text.

### mtDNAcn mediates the association between personality and mortality risk

The above preliminary observation that mtDNAcn was associated with mortality-related personality traits led us to a more detailed investigation on the relationship among personality, blood mtDNAcn, and mortality. In particular, we further extended our analysis to include four facets (*vulnerability, self-discipline, activity*, and *competence*) by creating a personality-mortality index (PMI) (see Methods for details) and performed a mediation analysis.

First, we observed an increasing mtDNAcn when moving from index zero (worst PMI) to index four (excellent PMI) in both study cohorts (Figure 3). Analyzing the personality-mortality index as a continuous variable showed that higher trait values were associated with higher mtDNAcn in both cohorts (Table 2). We confirmed the association between PMI and mortality by finding that the participants had significantly longer survival time with increasing PMI from zero to four (Figure 4).

**Figure 3.**
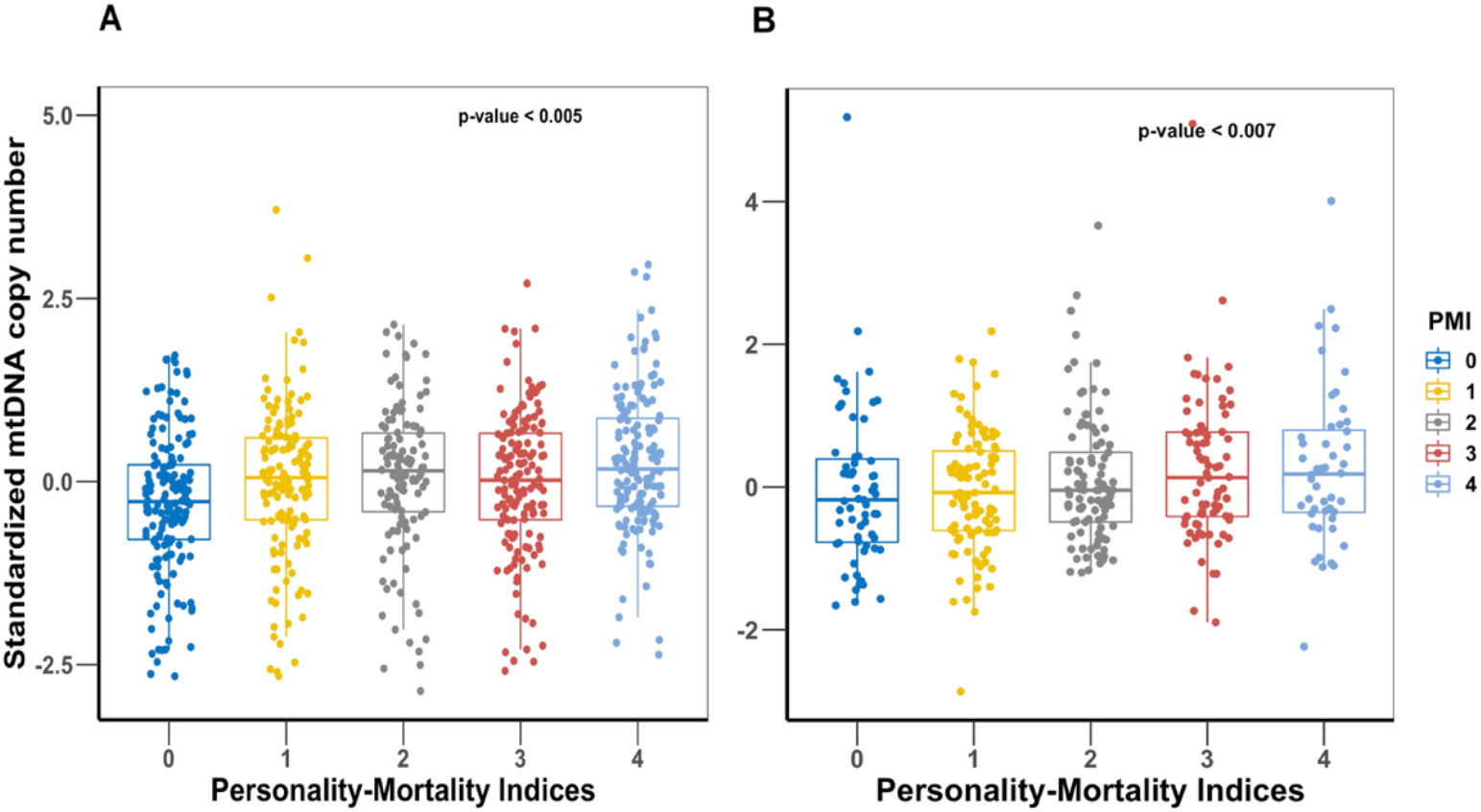
Comparison of mtDNAcn values amongst the five personality-mortality indices created from the *vulnerability* facet of Neuroticism, *activity* facet of Extraversion, *self-discipline* and *competence* facets of Conscientiousness. Panel A is BLSA cohort and panel B is SardiNIA cohort. The personality-mortality index with the lowest mortality risk is 4 and the index with the highest mortality risk is index 0. P-values of linear regression of mtDNAcn and the personality-mortality index are shown on the plots.

**Figure 4.**
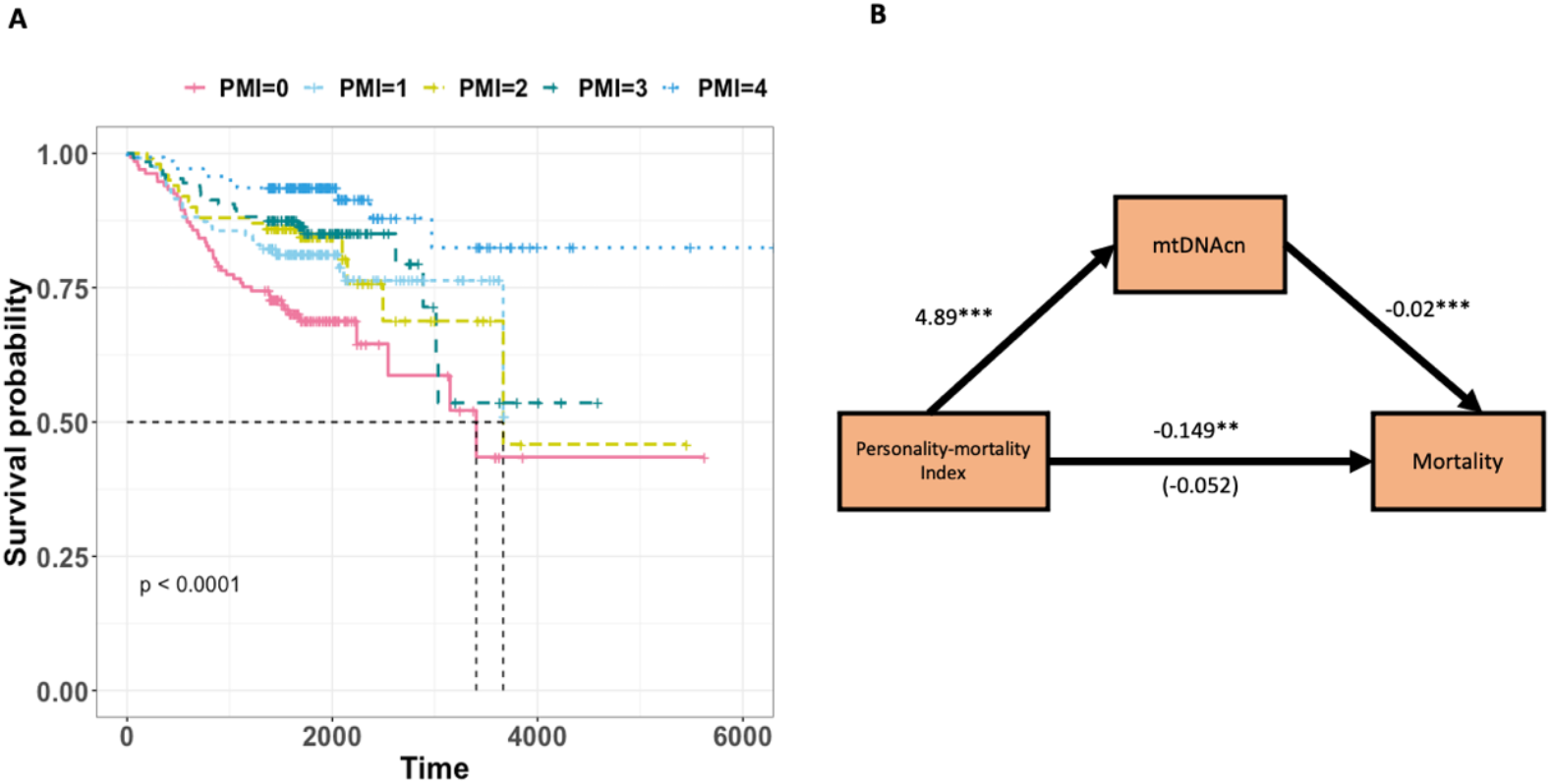
The effect of Personality-mortality Index (PMI) on mortality is fully mediated via mtDNAcn. Panel A is a Kaplan-Meier plot of time to death of BLSA participants in the five Personality Mortality Indices. Panel B shows the regression coefficients between PMI (predictor), mortality (outcome), and mtDNAcn (mediator) for the BLSA study. The effects of age and sex are adjusted for in the models. The indirect effect is (4.89) x (-0.02) = -0.097. We tested for the significance of the indirect effect using 5000 bootstrap samples and it was statistically significant (p value = 0.002). See Table 3 for more details.

Given the significant associations of mtDNAcn and specific personality traits with mortality, we then tested the hypothesis that the association between PMI and mortality may be mediated through mtDNAcn. We used structural equation modeling to test the indirect effect of PMI on mortality via mtDNAcn. The results are shown in Table 3 and Figure 4. We observed that for BLSA, PMI was a significant predictor of mortality. The indirect effect estimate showed that the effect of PMI on mortality was fully mediated via mtDNAcn. We tested the significance of the indirect effect using 5000 bootstrap samples, which yielded a significant p-value = 0.002 (estimate = -0.097). Similar results were obtained for SardiNIA, although they were not statistically significant, likely due to limited power because the Sardinian cohort is younger, and the mortality data are limited compared to the BLSA cohort. Also, the Sardinian analysis had reduced statistical power compared to the BLSA analysis because the personality measures are averaged across multiple visits for BLSA, which decreases the standard deviation (increasing the reliability) of effect estimates.

**Table 3.** Testing the direct and indirect effects of Personality-mortality index and mtDNAcn on mortality.

|  | **BLSA (n = 721)** | **SardiNIA (n = 395)** |
| --- | --- | --- |
| **Total Effect** | **Estimate (p value)** | **Estimate (p value)** |
| Personality-mortality index | -0.149 (0.001) | -0.022 (0.808) |
| **Full effects** |  |  |
| Personality-mortality index | -0.052 (0.260) | -0.019 (0.895) |
| mtDNAcn | -0.020 (0.000) | -0.001 (0.966) |
| **Indirect Effect** | **Estimate [Nonparametric Bootstrap 95% CI]** | **Estimate [Nonparametric Bootstrap 95% CI]** |
| mtDNAcn | -0.097 [-0.143; -0.03] | -0.004 [-0.169; 0.189] |

## Discussion

An extensive literature has reported the association of blood mtDNAcn with various psychological outcomes such as major depression (17, 18), autism (40), bipolar disorder (41), and stress (42– 44). Our study adds new information to this literature by reporting an association between mtDNAcn and personality traits. Our results show that higher mtDNAcn is significantly associated with a lower level of Neuroticism and higher levels of Extraversion, Openness, Agreeableness, and Conscientiousness in study participants. We demonstrate the robustness of these associations by showing that the direction and size of the effects are robust to adjustment for potential confounders such as age, sex, platelet count, and white blood cell parameters and are replicated across two cohorts. Consistent with Neuroticism, participants with more depressive symptoms had lower mtDNAcn than those without depressive symptoms. Finally, we report an effect of personality on mortality and demonstrate that this effect is partially mediated via mtDNAcn.

mtDNAcn is a biomarker of energetic health and cellular aging (16). The mechanism that controls the levels of mtDNAcn in tissues, blood, and plasma is not fully understood, and it is unclear whether the change of blood mtDNAcn reflects mitochondrial biogenesis, immune health, and/or hematopoiesis in the bone marrow (4). Increased mtDNAcn have been shown to have a different and potentially opposite indication depending on the underlying health condition (13). In population-based cohorts such as those involved in this study, increased mtDNAcn is generally positively correlated with mitochondria mass and thus is generally associated with higher oxidative capacity. Having a large pool of well-functioning mitochondria can positively influence health and generate signals that contribute to stress adaptation (16). Consistent with this interpretation, we found that the mtDNAcn levels were higher in younger participants, and our primary findings indicated that individuals with higher mtDNAcn levels had lower Neuroticism and were at lower risk of depression and lower risk of mortality. In adverse health conditions, however, it should be noted that increased mtDNAcn may not correlate with function (13, 45). Rather, it may indicate a compensatory upregulation of mtDNAcn (4), a phenomenon where mtDNAcn is increased to compensate for low energy cellular environment due to poor mitochondrial energetics (4, 15).

Personality traits influence behaviors, emotional states, and how individuals deal with stress. For example, personality traits are associated with how individuals assess stressful life circumstances (24) and what coping responses to the stressors (i.e., neuroendocrine responses, immune functioning and inflammation, cardiovascular responses) are elicited (46). Given these associations with stress and health-related behaviors, it is not surprising that personality traits are also related to better health, function, and longevity (27, 29, 34, 47–50). We observed a positive association between mtDNAcn and personality traits that are also related to health-relevant habits and conditions, such as non-smoking (51), healthy diet and obesity (52, 53), exercise and physical function (54), and preventive medical screenings (55). Thus, indirect pathways may exist to link personality and health-related behaviors to changes in mitochondrial biology. Our finding of an association between mtDNAcn and personality expands this broader literature on personality traits and health.

Reduced mtDNAcn may reflect reduced mitochondrial mass or loss of mtDNA due to mitochondrial dysfunction (4). Dysfunctional mitochondria that are not appropriately removed by mitophagy produce excess ROS, which causes oxidative stress. As a result of damage caused by oxidative stress, mitochondria release mitochondrial antigenic molecules called damage-associated molecular patterns (DAMPs) into the cytoplasm (16, 56). These released DAMPs contain mtDNA, which are unmethylated and thus act as antigenic fragments and elicit an innate immune response through toll-like receptors (57). This triggers an inflammatory response through cytokine release (16, 56). Prolonged inflammation, when unresolved, can lead to damage to cells and tissues and has been associated with a number of chronic diseases, including cardiovascular diseases (58), depression (59), and neurodegeneration (60). Our study shows that low mtDNAcn is associated with higher levels of Neuroticism and facets of Neuroticism, including depression. Of note, data from the SardiNIA sample indicates that Neuroticism is associated with higher interleukin-6 (IL-6) and C-reactive protein (CRP) (61). We observed a similar relationship between mtDNAcn and CES-D (depressive symptoms), and the effect becomes even more pronounced (almost doubling in effect size) when considering significant depressive symptoms (setting CES-D binary cutoff at 20 instead of at 16). Kim et al. (18) report a similar negative association between mtDNAcn and depression. However, other studies like Cai et al. (17) and Tyrka et al. (44) report a positive association of mtDNAcn with major depression, and several studies have found null associations (19–23). One explanation for these inconsistencies could be the heterogeneity of the depression phenotyping: the depression facet of the Neuroticism domain is a trait measure, and the CES-D measures symptoms, but neither is a diagnostic instrument for major depression. It should also be noted that although some past studies assayed for mtDNAcn from whole blood, they did not adjust for the effect of platelets, white blood cells, and other leukocytes subgroups on mtDNAcn as we do in this study. mtDNAcn estimates differ amongst the various blood cell types (62), and this significantly impacts whole-blood mtDNAcn estimates and should be adjusted for in association models (also see Discussion below).

mtDNAcn and personality are both predictors of mortality (25, 36, 63), thus making our finding of an association between the two even more intriguing. Understanding which variable affects the other in driving the effect on mortality is a significant step forward, but alternative models are similarly plausible. To test the hypothesis that mtDNAcn mediates the association between personality and mortality, we performed a causal mediation analysis. In this paper, we provide the first evidence of mtDNAcn significantly mediating the association between personality [including three facets capturing positive psychosocial states (activity, self-discipline, and competence) and an emotional stressor (vulnerability)] and mortality. Advances in mitochondrial research have presented the mitochondrion as a target of stress (64), and more recently, as a potential biological mediator of the stress-disease cascade (16). Picard and McEwen (16) provide a conceptual model for mitochondrial stress pathophysiology, outlining how mitochondria interact with psychosocial factors and stressors to influence health and lifespan outcomes. The underlying mechanism for this mediation is still an area of active research and is not fully understood, but the significance of our results remains clear: mtDNAcn is one mechanism in the pathway that connects personality and mortality.

This study also shows that mtDNAcn was negatively associated with age and male biological sex, which is consistent with previous reports (65). Also, higher mtDNAcn was significantly associated with higher levels of platelet count and percentages of lymphocytes, eosinophils, monocytes, and basophils. However, higher mtDNAcn was associated with lower white blood cell (WBC) count and lower neutrophil percentage. Neutrophils have relatively lower mtDNAcn compared to other leukocytes (62), and they form the largest proportion of WBC, which may explain the negative association of WBC count with mtDNAcn. A similar negative association between mtDNAcn and WBC count was observed by Liu et al. (66). The current study and other recent publications (67– 69) have highlighted the importance of adjusting for white blood cell parameters and platelet count when testing the association between traits and mtDNAcn measured in whole blood.

The primary strength of the current study is the use of two independent study cohorts, one in the US and the other in Europe: the top associations not only had significant p-values in both studies but also had very similar estimated effect sizes. Furthermore, in both cohorts, the associations of the personality traits and mtDNAcn ranked high when put in a broader context of mtDNAcn associations with other traits. The consistency of the results with the previous literature also clearly demonstrates the reproducibility of this current study. The way mtDNAcn is estimated using whole-genome sequence information from the two study cohorts is also seen as the gold standard going forward in this line of research (70), with major mitochondria consortia such as TopMed (66) employing the same approach. A limitation of the current study was the one-time assessment of mtDNAcn, which precluded a longitudinal analysis of changes in mtDNAcn. Personality was assessed only once in the SardiNIA cohort, but personality is relatively stable in adults over long periods (71, 72); therefore, not much information may be lost in a cross-sectional analysis of personality. Another drawback of this study is the difference in the distribution of age and mtDNAcn across the two cohorts. The differences in the average mtDNAcn between the two cohorts can be explained by the gap in the time at which sequencing was done; almost a decade passed between the time the two cohorts were sequenced. Despite these limitations, the associations between mtDNAcn and personality traits are clear and consistent across the two study cohorts.

In conclusion, we provide the first evidence of an association between mtDNAcn and personality. These results may provide further evidence of the link between mtDNAcn and psychosocial stress and suggest that blood mtDNAcn may correlate with changes in mitochondrial biology under stress. These initial findings will require further exploration to reveal the potential causal pathways between personality and mtDNAcn. We also provide novel insights into the effect of personality on mortality by showing that this effect is mediated through mtDNAcn.

## Materials and Methods

### Study cohorts’ description

#### Baltimore Longitudinal Study of Aging

The Baltimore Longitudinal Study of Aging (BLSA) is an ongoing scientific study that aims to understand risk factors and pathways that cause a decline in physical and cognitive function with aging. The study began in 1958 and is run by the National Institute on Aging Intramural Research Program and enrolls healthy volunteers. Blood samples were obtained from 955 individuals, and DNA was extracted from a red-cell-free buffy coat and sequenced on a HiSeq X Ten sequencer using 150 base-pair, paired-end cycles (version 2.5 chemistry, Illumina), as described elsewhere (73). For genetic homogeneity, we retained only participants with white ancestry for further analysis downstream. We further excluded participants who had developed Alzheimer’s Disease (AD) from further analysis because their NEO PI-R profiles are likely changed by clinical dementia (74). Therefore, 722 participants (mean age 75, ranging from 48 to 100, 48% were women) remained for downstream analysis. All participants gave written informed consent, with protocols approved by the Institutional Review Board of the Intramural Research Program of the National Institutes of Health.

#### SardiNIA Longitudinal Study

SardiNIA (39) is a longitudinal study of human aging and genetics of quantitative traits that has followed a genetically isolated population on the Italian island of Sardinia since 2000. The data used for this current study consist of 2,077 participants who had their DNA from whole blood sequenced using Illumina Genome Analyzer IIx and Illumina HiSeq 2000 instruments. We further excluded one of each pair of participants with a genetic relationship above the level of second-degree cousins. A total of 587 participants (mean age 57, ranging from 15 to 96, 62% women) remained for further downstream analysis. All participants gave written informed consent, with protocols approved by the Institutional Review Board of the Intramural Research Program of the National Institutes of Health.

#### Phenotype measures and definition

Personality was assessed using the **R**evised **NEO P**ersonality **I**nventory (NEO-PI-R) (26). The NEO-PI-R is an inventory of 240 items that examines an individual’s thoughts, feelings, and behaviors (26). Each item is answered on a five-point Likert scale ranging from “strongly disagree” to “strongly agree.” Responses to these items are scored to assess the *big five* personality traits (the five domains of personality traits): Neuroticism (N), Extraversion (E), Openness (O), Agreeableness (A), and Conscientiousness (C). In addition to the five domains, the NEO-PI-R also assesses six facets within each domain, for a total of 30 facets [for example, for the Neuroticism domain, the following six facets are assessed: Anxiety (N1), Angry Hostility (N2), Depression (N3), Self-Consciousness (N4), Impulsiveness (N5), Vulnerability (N6)]. Scores are standardized using a normative population to generate standardized *T scores* (mean =50, SD =10) (26). These *T scores* were used in the analysis.

BLSA participants completed the NEO-PI-R at each clinic visit (four assessment points on average), and we used the average across all assessment points for the downstream analysis. We did this because barring onset and progression of clinical dementia, which causes a drastic change in NEO PI-R profiles (49), NEO PI-R scores are quite stable in adults over long periods (71). Averaging the NEO-PI-R scores over multiple visits improves the BLSA analysis by reducing the standard deviation of the effects estimates and therefore increases statistical power. SardiNIA participants completed the NEO-PI-R once, and we used this one-time measure for downstream analysis. An extensive literature supports the validity and reliability of the NEO-PI-R scales in the United States and other countries, including Italy (75, 76). In both the BLSA and SardiNIA, the five traits have high internal consistency (alpha > .80), and in the BLSA, the test-retest stability of the five traits ranges from .78 to .83 over a follow-up of 10 years (72, 77). The correlations between the NEO PI-R traits are shown in Fig S5.

We also analyzed in both studies the measure of depressive symptoms using the Center for Epidemiologic Studies Depression Scale (CES-D) (78). The CES-D is a 20-item instrument assessing depressive mood and behavior over the past week. Responses to these statements are graded on a 4-point Likert scale ranging from “rarely or none of the time” to “most or all of the time.” The responses are scored from zero to three and summed across the 20 items to assess depressive symptoms (range 0 to 60). In general populations, a CES-D score of 16 is the clinical cutoff used to classify depressive symptoms (79, 80). However, among older populations, a cutoff score of 20 is used to improve the accuracy of the diagnosis of major depression (81, 82). We, therefore, analyzed two binary clinical cutoffs of CES-D (16 and 20 for significant depressive symptoms) and continuous scores in both studies. CES-D in both studies was assessed at a single time point.

#### mtDNA copy number estimation

The whole blood mitochondria DNA copy number (mtDNAcn) of each study participant was estimated from whole-genome sequence information using *fastMitoCalc* (83). This software implements the formula below, as described by Ding et al. (65).

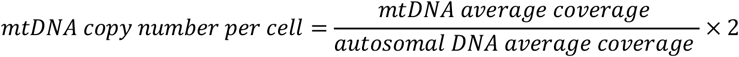

The above formula was derived from the fact that average sequencing coverage should be proportional to DNA copy number for autosomal DNA and mtDNA. Since there are two copies of autosomal DNA in each cell, the mtDNAcn is inferred as a ratio of average mtDNA coverage to autosomal DNA coverage multiplied by two. The average coverage of mtDNA and autosomal DNA is obtained using sequence alignment with *SAMtools* (84).

## Statistical Analysis

We performed initial simple linear regression of mtDNAcn values on variables such as age, sex, sequence coverage, platelet count, and white blood cell (WBC) parameters, including white blood cell count, percentages of the major leukocytes: lymphocytes, neutrophils, eosinophils, monocytes, and basophils. These initial assessments helped us determine which variables should be included as covariates in subsequent multiple linear regression models of mtDNAcn on NEO-PI-R traits.

As expected from previous findings reviewed in (4), age, sex, white blood cell count, platelet count, and percentages of neutrophils, lymphocytes, and basophils were significantly associated with mtDNAcn (*p*-value <0.0001). Sequence coverage and percentage of eosinophils were nominally associated with mtDNAcn (*p*-value <0.05), while percentage monocytes was marginally associated with mtDNAcn (*p*-value =0.0821). Therefore, all regression analyses relating mtDNAcn with personality traits or depression were adjusted for age, sex, white blood cell and platelet count, percent neutrophils, lymphocytes, basophils, and eosinophils, and sequence coverage.

Standardized values were used in regression analyses to make effect sizes more comparable across the two study cohorts. Therefore, all reported NEO-PI-R associations are expressed as the number of standard deviations change in mtDNAcn in response to higher NEO-PI-R trait values. A random-effect meta-analysis was used to obtain the combined estimates of effects and 95% confidence interval (CI) of each NEO PI-R/ CESD trait on mtDNAcn using the standardized effect sizes obtained from the two study cohorts.

### Personality, mtDNAcn, and mortality risk

In previous studies, the *vulnerability* facet of Neuroticism, the *activity* facet of *Extraversion*, and the *self-discipline* and *competence* facets of Conscientiousness were associated with mortality risk, with *vulnerability* (increased risk) and *self-discipline* (reduced risk) showing the strongest associations (25). To confirm these findings in the literature, we classified BLSA and SardiNIA participants according to *vulnerability* and *self-discipline* scores below and above the median level, and cross-classified participants into four groups by combinations of these two dichotomous variables (see Fig S6 for more details). We tested their association with mortality and mtDNAcn, using the low vulnerability/ high self-discipline group as a reference.

In subsequent analyses, we computed a personality-mortality index (PMI) using data from the four NEO PI-R facets (*vulnerability, activity, self-discipline*, and *competence*) that have been previously shown as significantly associated with mortality (25). For each of the four traits, we assigned a score of 0 or 1 to each individual based on the median value and the direction of the association of the trait with mortality. For example, individuals with *vulnerability* measures lower than the median received a score of 1 for the trait, and individuals with *self-discipline* measures higher than the median received a score of 1. The final PMI is the sum of the four scores for the four traits. The index ranged between 0 and 4, where a score of 0 means an individual has a high *vulnerability*, low *self-discipline*, low *activity*, and low *competence* (i.e., a poor PMI and potentially highest mortality risk). A score of 4 represents low *vulnerability*, high *self-discipline*, high *activity*, and high *competence*, indicating a favorable PMI and potentially lowest mortality risk. Scores of 1, 2, and 3 are various levels of improved PMI. We compared the mtDNAcn amongst the five indices (0, 1, 2, 3, and 4) of the PMI. We also tested for an association between PMI as a continuous variable and mtDNAcn, and death (a binary variable that classified study participants as dead or alive).

Finally, we used R package lavaan (version 0.6-7) to perform a mediation analysis to test whether mtDNAcn mediates the association between personality (i.e., PMI) and mortality risk. In general, there are three steps in a mediation analysis to show a potential mediator variable mediates the relationship between an independent variable and a dependent variable. Briefly, the three steps in our specific analysis are: 1) a total effect model, that regresses mortality (dependent variable) on PMI (independent variable) to confirm that PMI is a significant predictor of mortality; 2) a mediator model, that regresses mtDNAcn (mediator) on PMI to confirm that PMI is a significant predictor of mtDNAcn; 3) an outcome model, that regresses mortality on both mtDNAcn and PMI to confirm that a) mtDNAcn is a significant predictor of mortality, and b) the strength of the previously significant effect of PMI in Step 1) is now greatly reduced (i.e., the effect of PMI on mortality is largely indirect via mtDNAcn). In our analysis, the significance of indirect effect was estimated using 5,000 bootstrap samples.

## Data Availability

All data produced in the present study are available upon reasonable request to the authors

## Acknowledgments

Support for this work was provided by the Intramural Research Program of the National Institute on Aging (Z01-AG000693 and Z01-AG000949) and the National Institute of Neurological Disorders and Stroke of the National Institutes of Health. AT was also supported by the National Institute on Aging of the National Institutes of Health Grant R01AG068093.

## Supplementary Information Text

### Personality, mtDNAcn, and mortality risk

We studied the relationship between mtDNAcn and personality facets known to be associated with mortality risk. Supplementary Figure 6 compares the mtDNAcn amongst four distinct personality types (see Methods for details) created from the vulnerability facet of Neuroticism and self-discipline facet of Conscientiousness, which were the two individual facets most strongly associated with mortality. When comparing the mtDNAcn between the four personality types in the BLSA cohort, we observed that the personality type with the perceived highest mortality risk (HVLD: high vulnerability low self-discipline) had the lowest mtDNAcn amongst the four personality types. The type with a perceived lowest mortality risk (LVHD: low vulnerability high self-discipline) had a mean mtDNAcn value that is 0.23 standard deviations higher than the highest mortality type (HVLD) (p-value <0.0065). As a comparison of effect size relative to the slope of mtDNAcn and age after adjusting for all covariates, this difference in mtDNAcn between personality types corresponds to the difference in mtDNAcn observed across 0.6 years (about seven months) of life. A similar mtDNAcn difference was observed in the SardiNIA cohort: the mean mtDNAcn difference between the two personality types was 0.21 standard deviations. This difference corresponds to the difference in mtDNAcn observed across 0.3 years (a little over three months) of life.

**Fig. S1.**
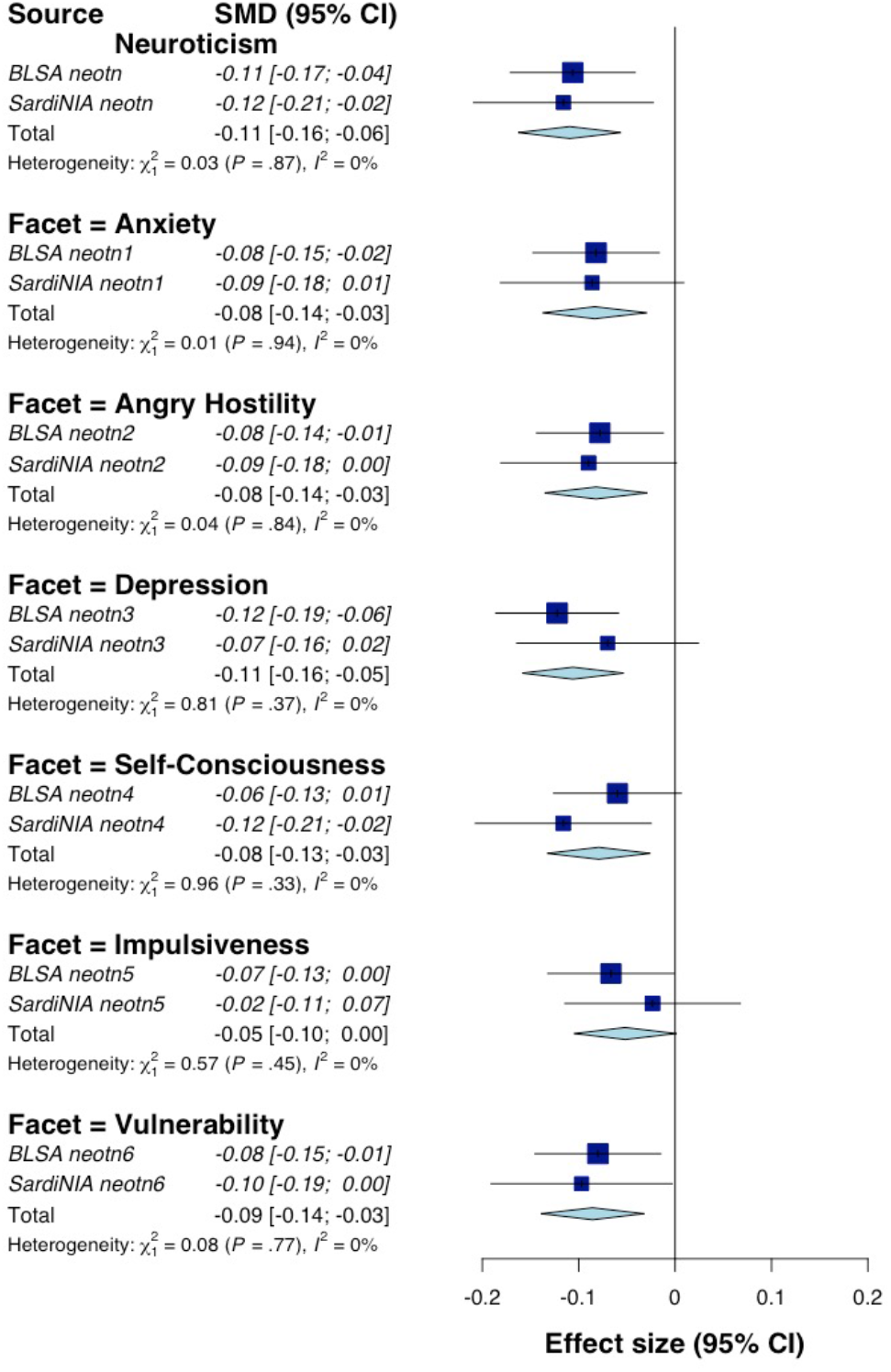
Random effect meta-analysis of Neuroticism domain of NEO PI-R.

**Fig. S2.**
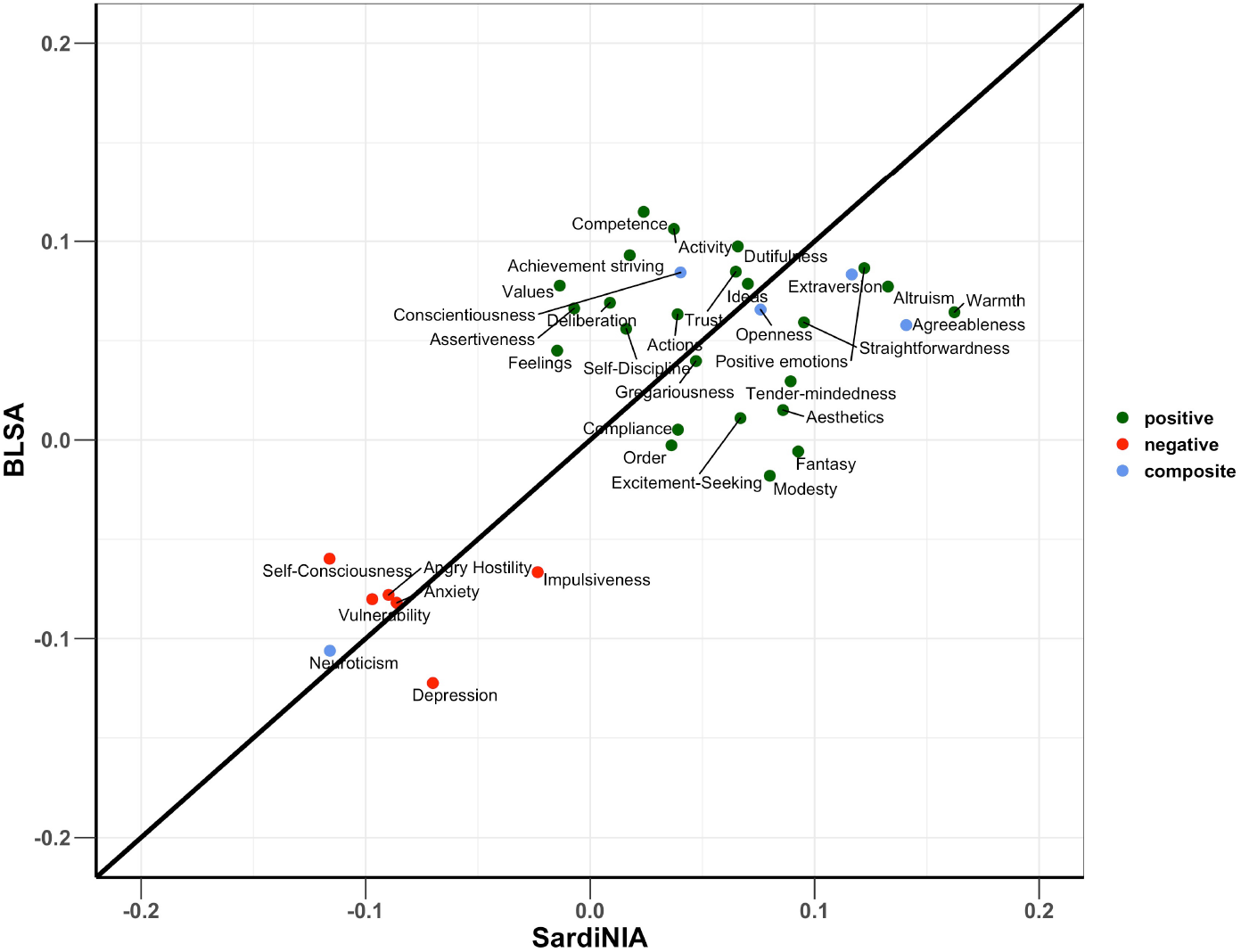
Comparison of effect sizes of NEO PI-R traits between the two cohorts. Regression model adjusted for the effects of age, sex, sequence coverage and white blood cell parameters. Color coding is green for positive personality types, red for negative personality types and blue for the composite scores for the Big five NEO PI-R domains.

**Fig. S3.**
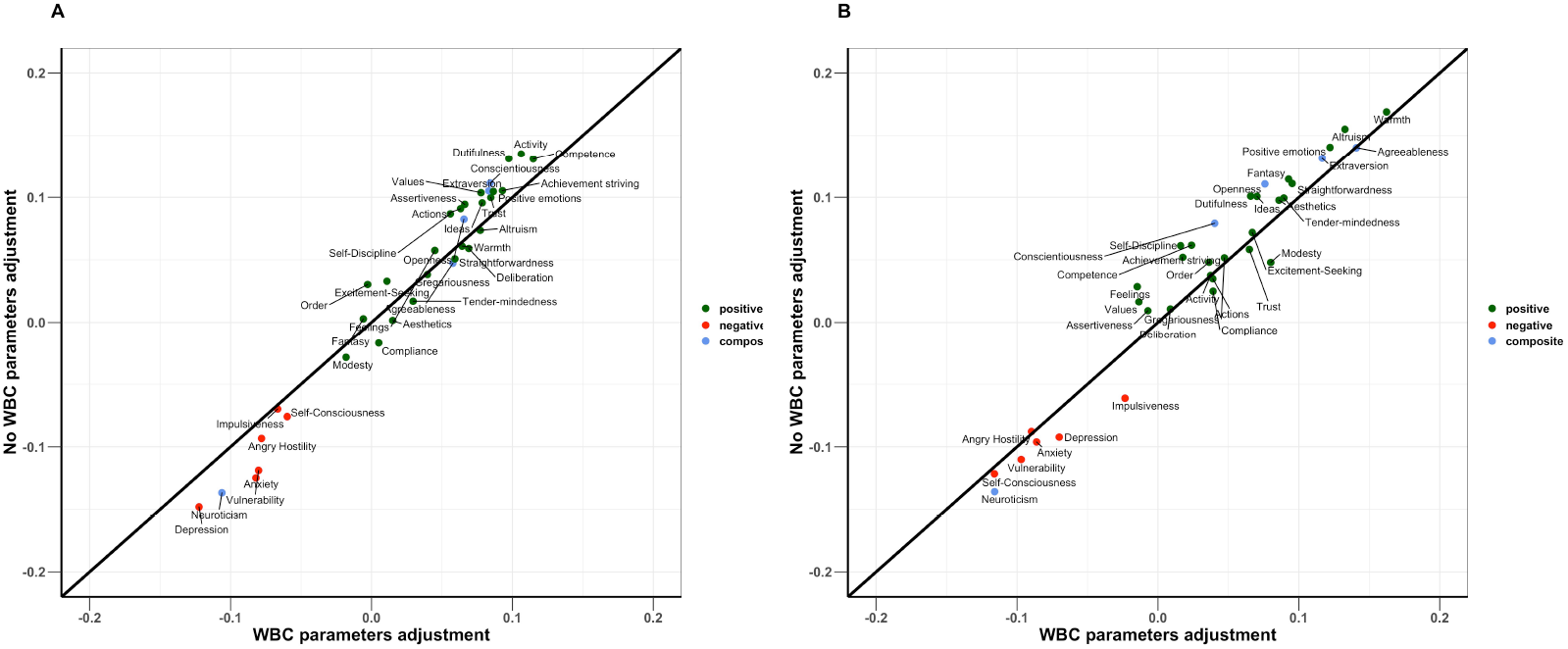
Comparison of effect sizes of NEO PI-R traits between models with WBC parameters adjustments and with no adjustments. Panel A is BLSA cohort and panel B is SardiNIA cohort. Color coding is green for positive personality types, red for negative personality types and blue for the composite scores for the Big five NEO PI-R domains.

**Fig. S4.**
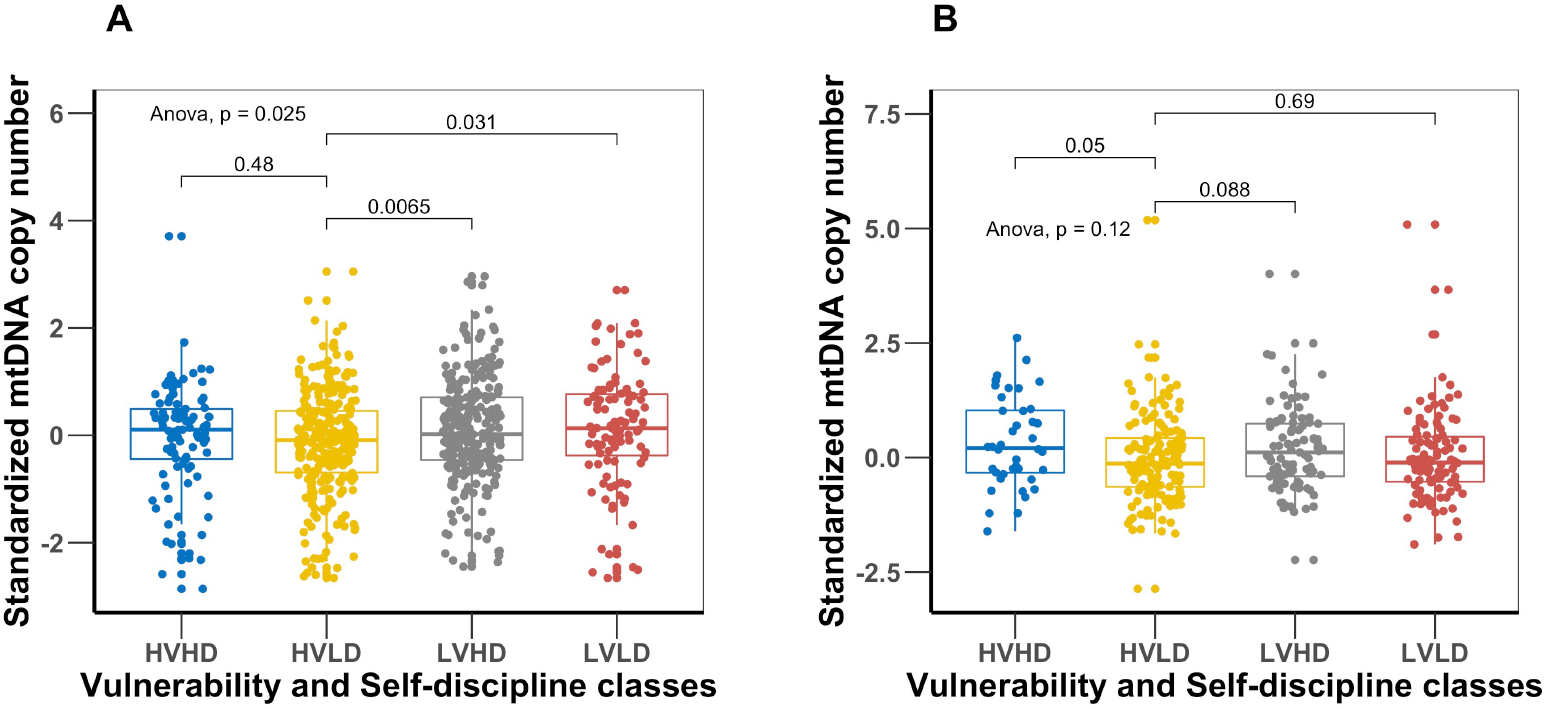
Comparison of mtDNAcn values amongst the four distinct personality types created from the vulnerability facet of Neuroticism and self-discipline facet of Conscientiousness. Panel A is BLSA cohort and panel B is SardiNIA cohort. The personality type with lowest mortality risk (LVHD: low vulnerability high self-discipline) has a significantly higher mean mtDNAcn value than the highest mortality risk type (HVLD: high vulnerability low self-discipline).

**Fig. S5.**
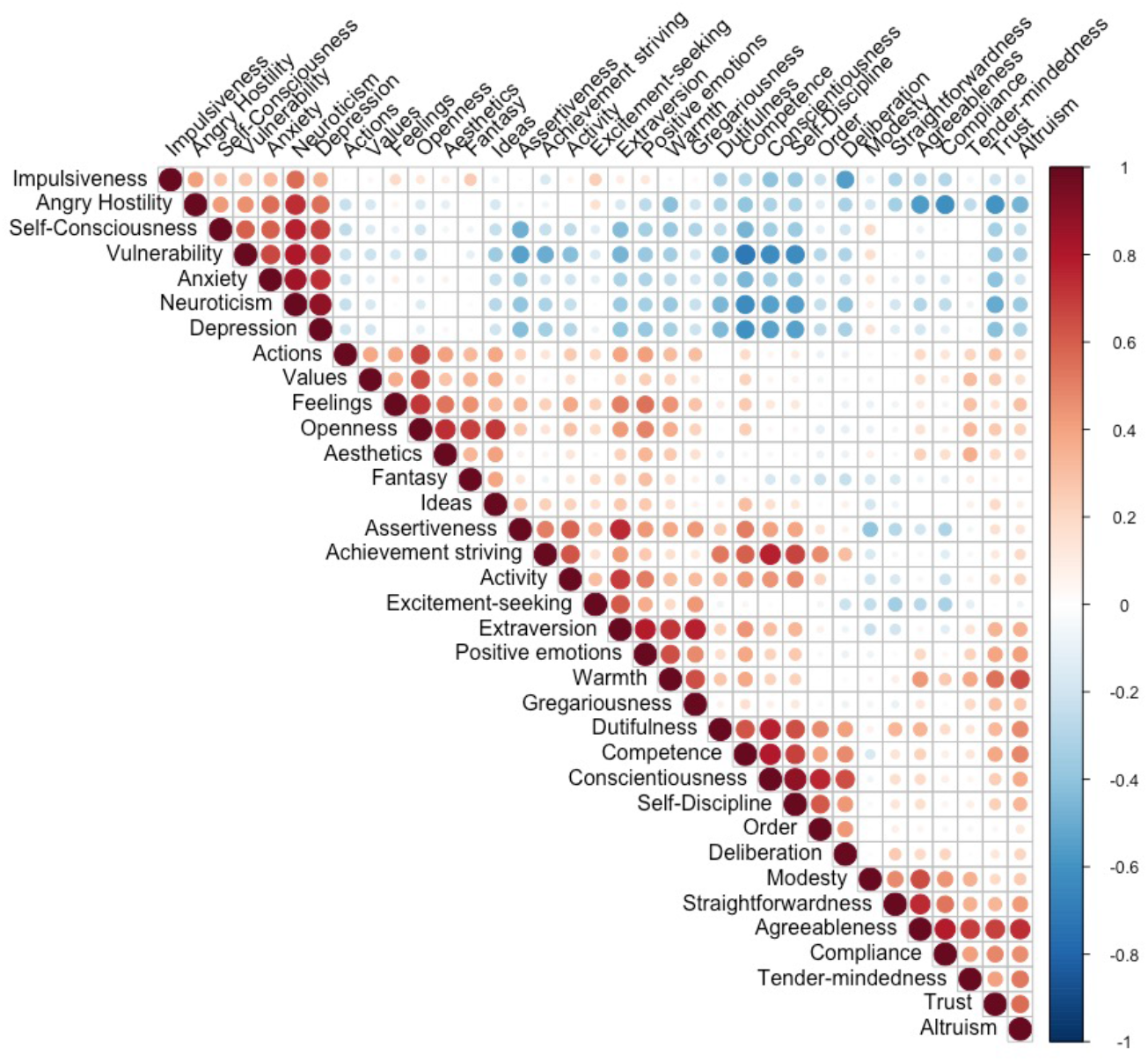
Correlation between NEO PI-R traits. The color intensity and size of circle are indication of size and significance of correlation.

**Fig. S6.**
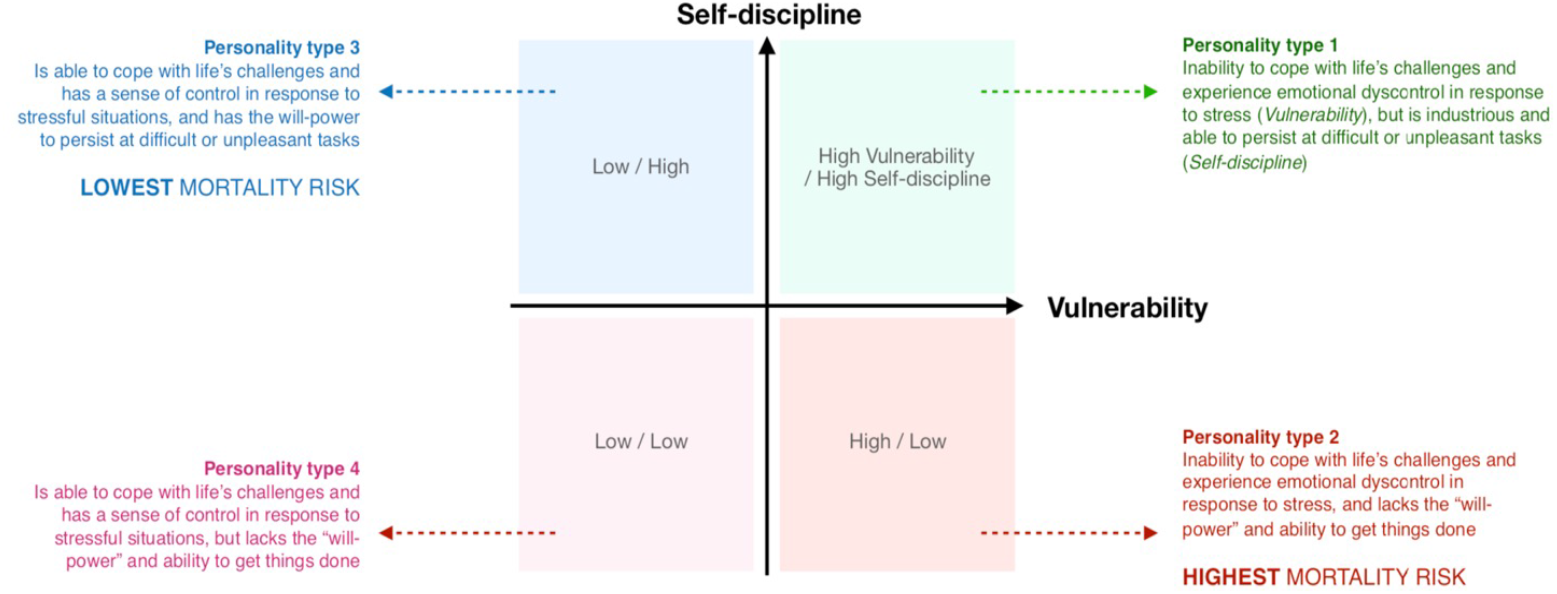
Four distinct personality types created from the Vulnerability facet of Neuroticism and Self-discipline facet of Conscientiousness.

**Table S1.** Association of age, sex, and white blood cell parameters with mtDNAcn.

| Trait | $\beta$ (p-value) of Model adjusted | | | |
| --- | --- | --- | --- | --- |
|  | No adjustment |  | Age, sex, coverage, and WBC parameters |  |
|  | BLSA | SardiNIA | BLSA | SardiNIA |
| Age | -0.025 (2.85e-13) | -0.0093 (0.003) | -0.014 (2.52e-5) | -0.0065 (0.028) |
| Sex | -0.61 (8.14e-17) | -0.25 (0.004) | -0.36 (1.21e-6) | -0.13 (0.11) |
| Platelet count | 0.004 (1.31e-7) | 0.0007 (0.30) | 0.003 (3.71e-6) | 0.001 (0.063) |
| WBC count | -0.13 (2.35e-10) | -0.14 (1.37e-8) | -0.097 (1.48e-6) | -0.08 (0.002) |
| Lymphocytes percent | 0.04 (5.5e-25) | 0.042 (5.19e-16) | -0.033 (0.23) | 0.073 (0.55) |
| Neutrophils percent | -0.041 (1.47e-28) | -0.041 (6.19e-18) | -0.063 (0.021) | 0.036 (0.77) |
| Eosinophils percent | 0.037 (0.031) | 0.043 (0.06) | -0.045 (0.16) | 0.076 (0.55) |
| Monocytes percent | 0.022 (0.082) | 0.062 (0.002) | -0.014 (0.57) | 0.082 (0.52) |
| Basophils percent | 0.44 (8.54e-5) | 0.11 (0.38) | 0.044 (0.69) | -0.076 (0.55) |

**Table S2.**
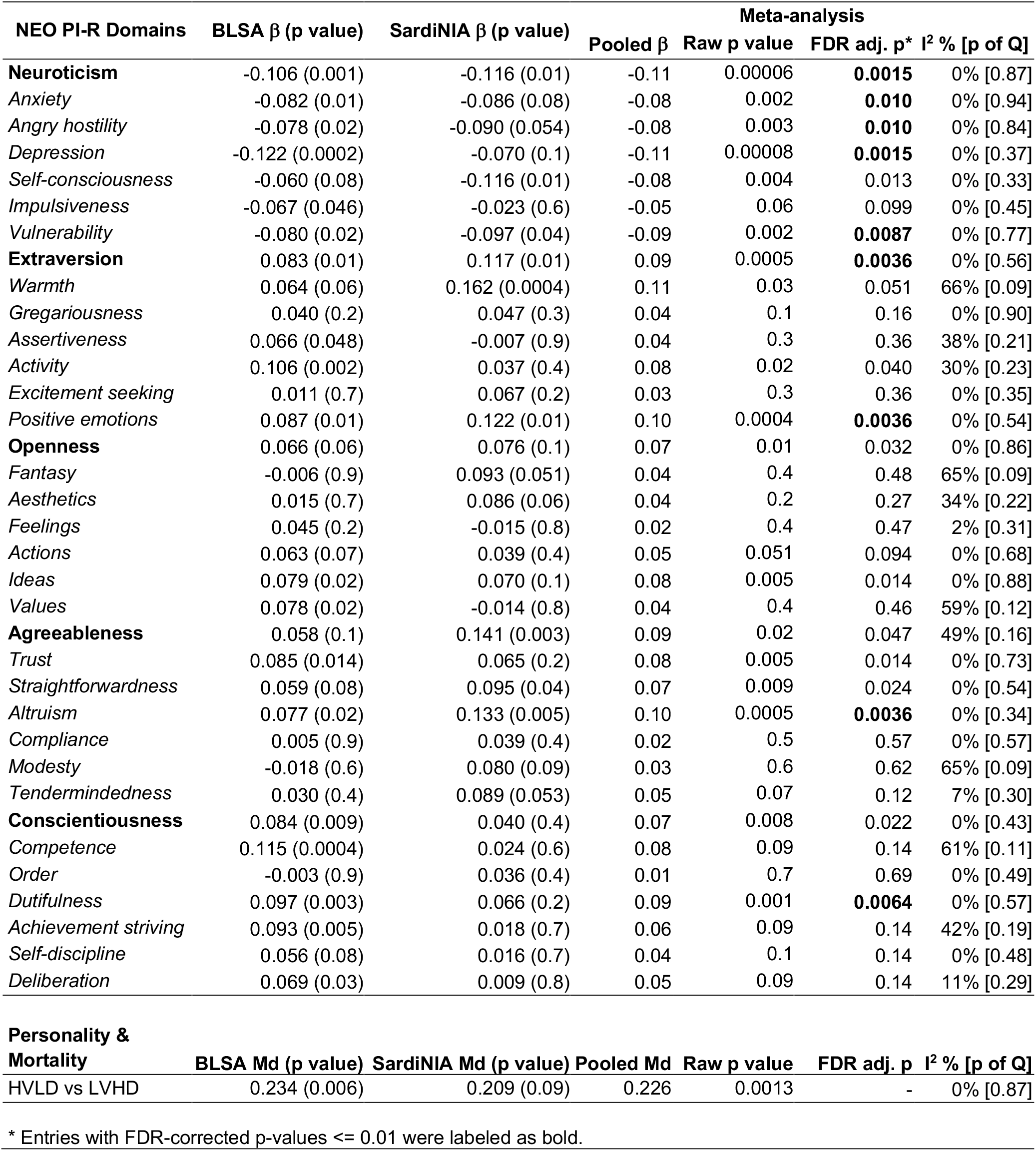
Association of NEO PI-R domains with mtDNAcn.

